# Development and validation of DNA Methylation scores in two European cohorts augment 10-year risk prediction of type 2 diabetes

**DOI:** 10.1101/2021.11.19.21266469

**Authors:** Yipeng Cheng, Danni A Gadd, Christian Gieger, Karla Monterrubio-Gómez, Yufei Zhang, Imrich Berta, Michael J Stam, Natalia Szlachetka, Evgenii Lobzaev, Nicola Wrobel, Lee Murphy, Archie Campbell, Cliff Nangle, Rosie M Walker, Chloe Fawns-Ritchie, Annette Peters, Wolfgang Rathmann, David J Porteous, Kathryn L Evans, Andrew M McIntosh, Timothy I Cannings, Melanie Waldenberger, Andrea Ganna, Daniel L McCartney, Catalina A Vallejos, Riccardo E Marioni

## Abstract

Type 2 diabetes mellitus (T2D) presents a major health and economic burden that could be alleviated with improved early prediction and intervention. While standard risk factors have shown good predictive performance, we show that the use of blood-based DNA methylation information leads to a significant improvement in the prediction of 10-year T2D incidence risk.

Previous studies have been largely constrained by linear assumptions, the use of CpGs one-at-a-time, and binary outcomes. We present a flexible approach (via an R package, *MethylPipeR*) based on a range of linear and tree-ensemble models that incorporate time-to-event data for prediction. Using the Generation Scotland cohort (training set n_cases_=374, n_controls_=9,461; test set n_cases_=252, n_controls_=4,526) our best-performing model (Area Under the Curve (AUC)=0.872, Precision Recall AUC (PRAUC)=0.302) showed notable improvement in 10-year onset prediction beyond standard risk factors (AUC=0.839, PRAUC=0.227). Replication was observed in the German-based KORA study (n=1,451, n_cases_ = 142, p=1.6×10^-5^).

## Introduction

Diabetes mellitus is one of the most prevalent diseases in the world and a leading cause of mortality. Around half a billion people live with diabetes worldwide, with type 2 diabetes (T2D) making up about 90% of these cases [1]. Individuals with diabetes can suffer from debilitating complications including nerve damage, kidney disease and blindness [2]. The disease also increases the future risk of dementia and cardiovascular disease [3], with recent studies highlighting obesity and T2D as risk factors for COVID-19 disease severity and ICU admission [4]. Furthermore, risk of complications increases over time and is exacerbated if blood-glucose levels are poorly managed. Despite developments in the way T2D can be managed for patients, these treatments are reactive, focusing on patients that have already been diagnosed. Early intervention with metformin or lifestyle changes have been shown to delay onset of T2D, although they did not reduce the risk of all-cause mortality [5].

Beyond public health costs, T2D also presents a substantial financial burden to the NHS, with estimated annual spending of £10 billion on diabetes in the UK. Around 80% of these costs are for treatment of complications, many of which are preventable with early intervention [6].

While the mechanisms of insulin resistance in T2D are well-known, the interactions between genetic and environmental factors that increase T2D susceptibility are less understood. Previous T2D risk prediction models have used a range of health risk factors [7]. However, these have not utilised epigenetic information. Epigenetics is the study of heritable changes to DNA that do not modify its nucleotide sequence. A commonly studied form of this is DNA methylation (DNAm), whereby methyl groups are attached to the DNA molecule - most commonly to the 5-carbon on a cytosine in a cytosine-guanine pair (CpG). Due to its involvement with gene expression and gene-environment interactions, DNAm can provide dynamic predictive information for disease risk for an individual. For example, Epigenetic Scores (EpiScores) built via penalised regression models have been used to show that weighted linear CpG predictors can explain a substantial proportion of phenotypic variance (R^2^) of modifiable health factors including body mass index (BMI) (12.5%), HDL cholesterol (15.6%) and smoking status (60.9%) [8]. Blood-based DNAm is of particular interest in predictive modelling and biomarker development due to its comparatively non-invasive sampling procedure. EpiScores have also shown the ability to explain up to 58% of variance in plasma protein levels are associated with a number of incident diseases including T2D and several comorbidities [9]. Epigenome-wide association studies (EWAS) have identified a number of CpG sites significantly associated with T2D [10–14] as well as related risk factors such as cardiovascular disease [15] and obesity [16, 17]. While these provide some predictive performance for T2D prevalence, incident T2D has been less well studied. One such EWAS with 563 cases and 701 controls identified 18 CpGs associated with incident T2D but did not consider any prediction models [10]. Given that preventative lifestyle changes have been shown to effectively reduce T2D onset [18], prediction of T2D incidence years ahead of time would be greatly beneficial in stratifying populations so those at high risk can be monitored and treated with early interventions.

Currently, most studies generating DNAm predictors consider marginal CpG effects or assume only linear additive effects between CpGs. The use of predictive models that can incorporate both interaction and non-linear effects could capture more complex relationships between variables, resulting in greater prediction accuracy. Therefore our study aims to evaluate both the additional predictive benefit that DNAm can provide for 10-year T2D risk and the applicability of linear and tree-ensemble survival models.

Here, we use one of the world’s largest studies with paired genome-wide DNAm and data linkage to electronic health records (EHR), Generation Scotland (n=14,613, n=626 incident T2D cases over 15 years of follow-up), to develop and validate epigenetic EpiScores for T2D. We show the added contribution of these EpiScores to prediction over and above standard risk factors e.g. age, sex and BMI and externally validate these results in the KORA S4 cohort.

## Methods

### Reporting of Analysis Pipeline and Results

To enhance reproducibility, the analysis pipeline and results presented in this study have been reported via the TRIPOD checklist [19] (**Supplementary File 1**).

### Generation Scotland

Blood-based DNAm and linked health data were obtained from Generation Scotland [20], a family-structured population-based cohort. The cohort consists of 23,960 volunteers across Scotland aged 18-99 years at recruitment (between 2006 and 2011), of whom over 18,000 currently have genome-wide DNAm data available (Illumina EPIC array). In DNAm quality control, CpG sites were filtered by removing those with low bead count in ≥5% of samples or a high detection p-value (>0.05) in more than 5% of samples. Probes on X and Y chromosomes were also removed. Samples were filtered by removing those with a mismatch between predicted and recorded sex or ≥ 1% of CpGs with detection p-value > 0.05. Missing CpG values were mean-imputed. To enable the predictors to be applied to existing cohort studies with older Illumina array data, CpGs were filtered to the intersection of the 450k and EPIC array sites (n=453,093 CpGs).

This study considered DNAm data from three large subsets of the GS cohort, with 5,087 (Set 1), 4,450 (Set 2) and 8,877 (Set 3) individuals. Processing took place in 2017, 2019 and 2021 respectively. Set 1 and Set 3 included related individuals within and between each set while all individuals in Set 2 were unrelated to each other and to individuals in Set 1 (genetic relationship matrix (GRM) threshold <0.05). In our experiments, the training set consisted of Sets 2 and 3 combined and Set 1 was used as the test set. To avoid the presence of families with individuals across both the training and test set, any individuals in the training set from the same family as an individual in the test set were excluded from the analysis (n_excluded_=3,138).

Participant health measures including age, BMI, sex, self-reported hypertension and family (parent or sibling) history of T2D were taken at baseline (DNAm sampling) via questionnaire. BMI was calculated as the individual’s weight in kilograms divided by the square of their height in metres. Missing values in the Set 1 health measures were treated as missing-completely-at-random and the corresponding individuals were excluded (n_Set 1_=99). This was not performed in Sets 2 and 3 as the health measures were used for incremental modelling (Set 1 only).

Disease cases were ascertained through data linkage to NHS Scotland health records consisting of hospital (ICD-10 codes) and GP records (Read2 codes). Prevalent cases were identified from a baseline questionnaire (self-reported) or from ICD-10/Read2 codes dated prior to baseline and removed from the dataset. Type 1 and juvenile cases were treated as control observations. A total of 757 incident cases were observed over the follow up period (from recruitment date to 01/2022) and after preprocessing, 626 cases remained. Mean time-to-T2D-onset was 5.9, 5.4 and 6.0 years for Sets 1, 2 and 3 respectively, with ranges of 0.2 – 14.8 (Set 1), 0.2 – 14.8 (Set 2) and 0.1-14.8 (Set 3) years. In GP record-derived cases, 81% of cases had a C10F. “type 2 diabetes mellitus” code; 12% had a C10.. “diabetes mellitus” code and 4% had a C109. “Non-insulin dependent diabetes mellitus” code. The full list of included and excluded terms are given in **Supplementary Table 1.**

### Composite Protein EpiScore

A composite protein EpiScore model for incident T2D was trained using a set of 109 protein EpiScores previously shown to associate with a range of incident diseases [9]. For each protein, the EpiScore was calculated for each subject in the training and test sets. A Cox proportional-hazards (Cox PH) lasso model was fit to the training set with the 109 protein EpiScores (scaled within set to mean of 0 and variance 1) as features. The linear predictor from the Cox PH lasso model was then used as the composite protein EpiScore in the incremental test set model.

### Direct EpiScore

The direct EpiScore Cox PH lasso model for incident T2D was fit to the DNAm data in the training set. Due to memory limitations in the model fitting R package (glmnet [21]), the CpGs were filtered to the 200,000 sites with highest variance. The linear predictor from the Cox PH lasso model was then used as the direct EpiScore in the incremental test set modelling. For tree ensemble models, the Cox PH lasso-selected CpGs were used as input and the 10-year onset risk was subsequently used as the direct EpiScore.

### Outcome Definition for 10-Year Onset Prediction

Linkage to NHS Scotland health records provided dates for disease diagnoses from which age-at-onset was calculated. Along with age at baseline (DNAm sampling), these were used to calculate the time-to-event, measured in years, for each individual. For incident T2D cases and controls, time-to-event was defined as the time from baseline to disease onset and censoring, respectively. Controls were censored at the latest date of available GP records (09/2020).

While all models were trained as survival models, our primary prediction outcome was incident T2D diagnosis within 10 years. Therefore, predictions on the test set were calculated using the 10 year onset probability (one minus survival probability). When calculating binary outcome metrics, cases with time-to-event (TTE) > 10 were treated as controls. These metrics included confusion matrices, area under the receiver operating characteristics curve (AUC) and area under the precision-recall curve (PRAUC). After preprocessing and case thresholding (TTE > 10), there were 218 cases and 4,560 controls in the test set.

The numbers of individuals/cases and controls after each preprocessing step are shown in Supplementary Figure 1.

### Incremental Modelling

Composite protein EpiScores and direct EpiScores were generated in the training dataset using different machine learning methods, via the *MethylPipeR* package (Figure 1), before being applied to the test set in an incremental modelling approach (further detail in **Supplementary Methods**). In the test set, a (null) risk factors-only model was defined via a Cox PH model for T2D with age, sex, BMI, self-reported hypertension, and self-reported family (sibling or parental) history of diabetes as predictors. A multitude of risk factors have been used in previous T2D risk prediction tools, a majority of which include the set that we have used in this study. While additional risk factors, such as waist-hip ratio, may also be relevant [7], we selected the null model covariates based on those used in the Diabetes UK type 2 diabetes ‘Know Your Risk’ tool to compare our results to an existing widely utilised tool. This was with the exception of ethnicity, due to the relative homogeneity of the GS cohort. These also closely match the top risk factors identified in a systematic review of previous T2D risk predictors (see Figure 2 in ref. [7]).

**Figure 1.**
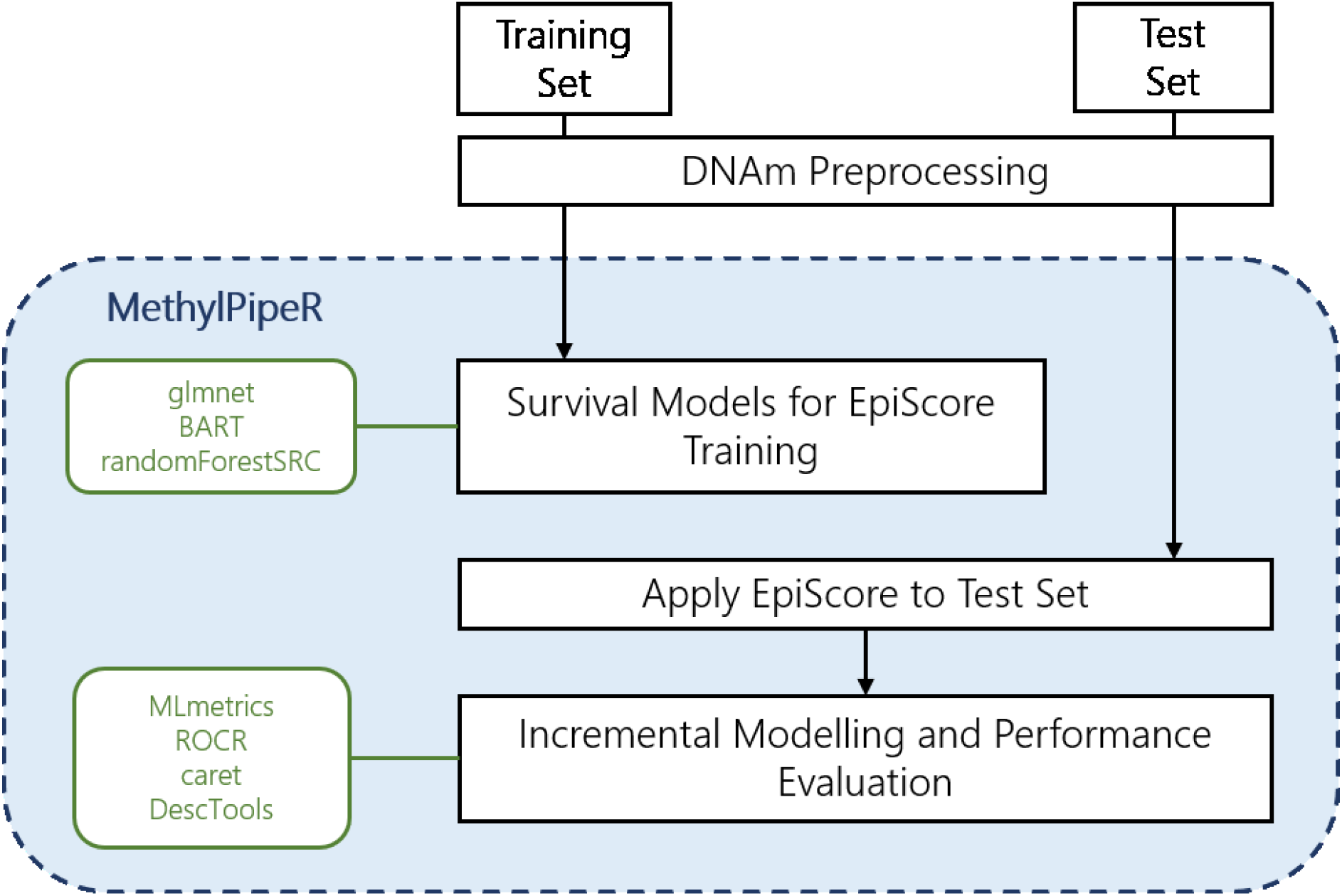
The prediction pipeline and functionality provided in *MethylPipeR*

**Figure 2.**
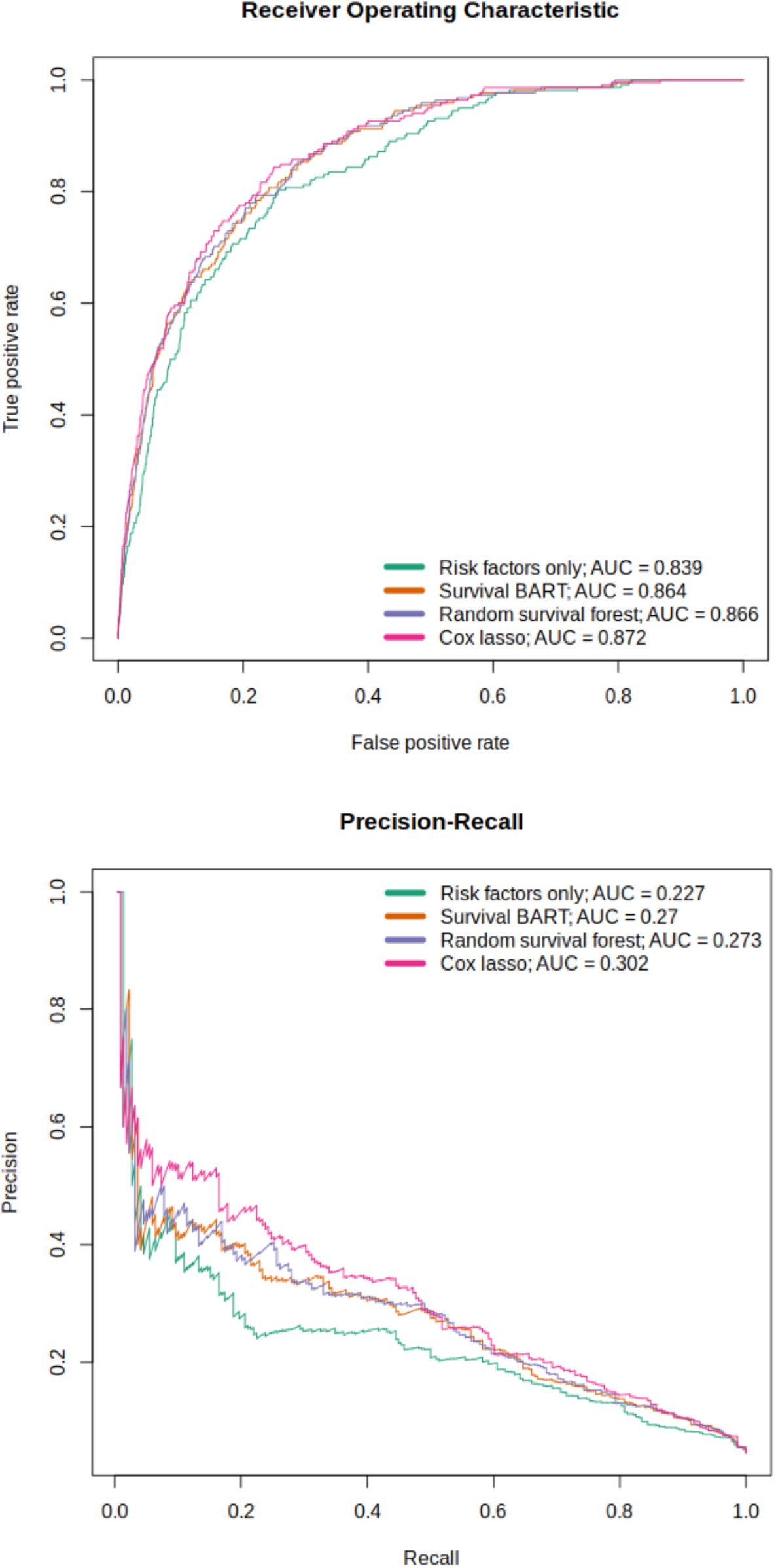
ROC and PR curves for the full models with Cox PH Lasso, Random Survival Forest and Survival BART direct EpiScores.

### Penalised Regression Predictors

Since the number of CpGs (n_CpG_=200,000) was much greater than the number of rows in the training set (n=9,835 after preprocessing), a regularisation method was required to reduce overfitting of the Cox PH regression models.

Penalised regression models reduce overfitting by applying a regularisation penalty in the model fitting process. This forces regression parameters to remain small, or possibly to shrink them to zero. The latter allows the method to be used for variable selection, by keeping only the variables with resulting non-zero coefficients.

Lasso penalisation was fit to the training set DNAm and protein EpiScores using glmnet [21, 22] via *MethylPipeR* with the best shrinkage parameter (λ) chosen by 9-fold cross-validation.

### Tree Ensemble Models

Tree ensembles are non-parametric models capable of estimating complex functions using a set of decision trees. Two tree ensemble approaches were used: random survival forest (RSF) [23] and survival Bayesian Additive Regression Trees (sBART) [24]. Random forest [25] is an ensemble machine learning model that estimates a function by averaging the output from a set of independently trained decision trees. During model fitting, each tree is built using a different subset of the variables from the training set to prevent individual trees from overfitting to the whole dataset. sBART is a non-parametric method that estimates a function as a sum over a set of regression trees. sBART incorporates the ability to model both additive and interaction effects and has shown high predictive performance in comparison with similar methods [24, 26].

RSF and sBART were fit to the training set using R packages randomForestSRC (version 2.11.0) [27] and BART (version 2.9) [28] respectively, via *MethylPipeR*. Details on hyperparameter selection are given in **Supplementary Methods**.

Due to computational limitations and probable overfitting in using the tree ensemble models on all CpGs in the dataset, variable pre-selection was based on the coefficients in the penalised Cox PH models. Each tree-ensemble model was evaluated with the features corresponding to non-zero coefficients from the Cox PH lasso model.

### Evaluating Predictive Performance

Survival models can be used to predict the risk of incident T2D for an arbitrary prediction period. Here we focus on classification performance for the binary outcome defined by 10-year T2D incidence. EpiScores were calculated as one minus 10-year survival probabilities and the binary outcomes were calculated by truncating observed TTE at 10-years (see *Outcome Definition for 10-Year Onset Prediction)*.

AUC and PRAUC were calculated as measures of predictive performance as the discrimination threshold was varied. PRAUC is more informative in situations where there is a class imbalance in the test set [29]. Additionally, binary classification metrics consisting of sensitivity (recall), specificity, positive predictive value (PPV/precision) and negative predictive value (NPV) were calculated. These metrics require selection of a discrimination threshold to assign positive/negative class predictions and have varying behaviour as this threshold is altered. Therefore, each of the metrics were calculated at a range of thresholds between 0-1 in increments of 0.1.

Model calibration was examined by comparing predicted probabilities with actual case/control proportions. The test data was sorted by predicted probability and divided into bins; the mean predicted probability and the proportion of cases was calculated for each bin.

### Selected-CpG Comparison with EWAS Catalog

The CpG sites selected by the Cox PH lasso were queried in the EWAS Catalog [30] to identify traits that have previously been linked to these sites. The EWAS catalog is a database allowing users to search EWAS results from existing literature. We performed a tissue-agnostic query using the selected CpGs and filtered results to those with an epigenome-wide significance threshold of P < 3.6 x 10^-8^ in studies with a sample size > 1,000 [31]. Almost all (739 out of 742; 99.6%) of the post-filter results were from blood-based studies. The remaining results were from saliva and prefrontal cortex-based studies.

### Validation in KORA S4

The Cox PH lasso model using the direct EpiScore was applied to the KORA S4 cohort [32]. This cohort consisted of 1,451 individuals in southern Germany, aged 25-74 years. Cohort summary details are shown in **Supplementary Table 2.** Individuals with missing values in the health measures were removed from the dataset. Missing CpG values in the DNAm data were mean-imputed. Similar to the approach in the Generation Scotland test set, an EpiScore was computed for each individual in the KORA dataset. Evaluation was then performed using an incremental modelling approach. Additional cohort and methods details (such as the outcome definition, follow-up period and preprocessing numbers) are provided in **Supplementary methods**.

### EpiScore Prediction of Ongoing Symptomatic COVID-19/Hospitalisation

The subset of the Generation Scotland cohort with reported COVID-19 infection (clinically-diagnosed or positive test from linked test data), who had also participated in the CovidLife study [33] were used for prediction of ongoing symptomatic COVID-19 and hospitalisation from COVID-19 (n=713). Ongoing symptomatic COVID-19 cases were defined here as individuals with self-reported symptoms lasting ≥ 4 weeks [34]. Hospitalisation cases were defined as hospital admissions with accompanying ICD10 codes U07.1 (confirmed COVID-19 test) and U07.2 (clinically diagnosed), derived from the Scottish Morbidity Records (SMR01). Details of the method and summary statistics are shown in **Supplementary methods** and **Supplementary Table 3.**

## Results

After preprocessing, the mean time-to-onset of T2D was 5.7 and 5.9 years for the training (n=374 cases) and test (n=252 cases) sets, respectively. Mean age-at-onset was also similar between the training and test set at 61.2 and 60.4 years and the mean BMI for cases (at baseline) was 31.7 and 32.2 kg/m^2^. The full set of cohort summary details for cases and controls in both sets are shown in Table 1. The machine learning prediction pipeline of the *MethylPipeR* package is shown in Figure 1.

**Table 1.**
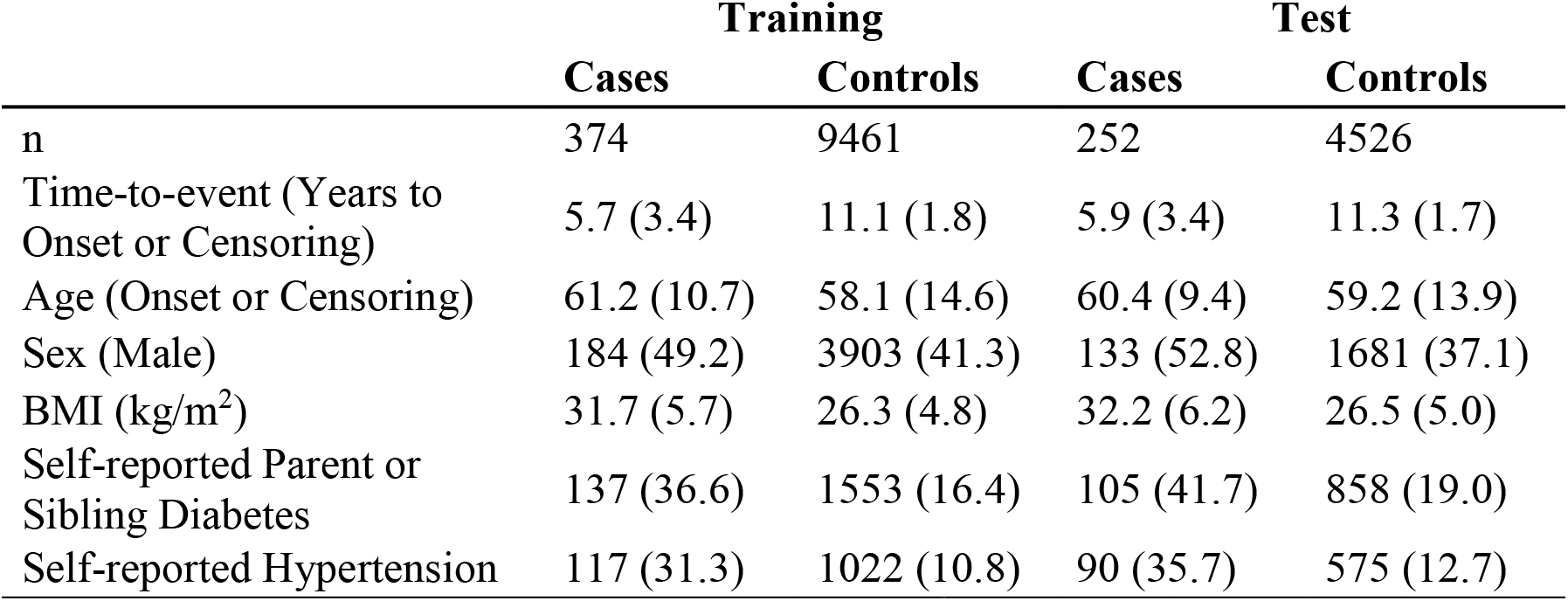
Summary information for the Generation Scotland training and test sets. Summary information is mean (SD) or n (%).

|  | **Training** |  | **Test** |  |
| --- | --- | --- | --- | --- |
|  | **Cases** | **Controls** | **Cases** | **Controls** |
| n | 374 | 9461 | 252 | 4526 |
| Time-to-event (Years to Onset or Censoring) | 5.7 (3.4) | 11.1 (1.8) | 5.9 (3.4) | 11.3 (1.7) |
| Age (Onset or Censoring) | 61.2 (10.7) | 58.1 (14.6) | 60.4 (9.4) | 59.2 (13.9) |
| Sex (Male) | 184 (49.2) | 3903 (41.3) | 133 (52.8) | 1681 (37.1) |
| BMI (kg/m<sup>2</sup>) | 31.7 (5.7) | 26.3 (4.8) | 32.2 (6.2) | 26.5 (5.0) |
| Self-reported Parent or Sibling Diabetes | 137 (36.6) | 1553 (16.4) | 105 (41.7) | 858 (19.0) |
| Self-reported Hypertension | 117 (31.3) | 1022 (10.8) | 90 (35.7) | 575 (12.7) |

### Null Model for the Incremental Modelling Approach

A Cox PH model in the test set with age, sex, BMI, self-reported hypertension, and family history of diabetes as predictors yielded good classification metrics: AUC=0.839, PRAUC=0.227.

### Incremental model using Direct EpiScore derived using Cox PH lasso, RSF and sBART

In the risk factors + direct EpiScore test set model, Cox PH lasso performed the best, showing an AUC and PRAUC of 0.870 and 0.299, respectively (p=3.6×10^-27^ for the EpiScore coefficient). This corresponds to an increase of 3.1% and 7.2% over the standard risk factors model.

Overall, the tree-ensemble models used for the direct EpiScore resulted in lower performance compared to Cox PH lasso. AUC values for RSF and sBART were 0.852 and 0.840 and PRAUC values were 0.247 and 0.230, respectively (**Supplementary Table 4**). P-values for the EpiScore coefficients and ROC curves for all models are given in (**Supplementary Table 5**).

### Incremental model using Composite Protein EpiScore derived using Cox PH lasso

The composite protein EpiScore showed good performance with AUC and PRAUC of 0.864 and 0.270, respectively (EpiScore coefficient p=1.61×10^-18^). The increase in PRAUC was smaller for the composite protein EpiScore compared to the direct EpiScore but still a notable improvement over using risk factors only.

### Incremental model using Composite Protein and Direct EpiScores

The full model (risk factors + composite protein EpiScore + direct EpiScore) with a Cox PH lasso direct EpiScore gave an AUC and PRAUC of 0.872 and 0.302 respectively. The full models with RSF- and sBART-derived direct EpiScores showed AUCs of 0.866 and 0.864, respectively. The corresponding PRAUC values were 0.273 and 0.270. Therefore, the best overall model used direct and composite protein EpiScores from Cox PH lasso models. The ROC and PR curves for the full models and risk factor only model are shown in Figure 2.

### Binary Classification Metrics and Model Calibration

**Supplementary Table 6** shows how confusion matrix metrics vary for the null (risk factors only) model and the Cox PH lasso model across a range of probability classification thresholds. As expected, as the classification probability threshold is increased, sensitivity and NPV decrease while specificity increases. The effects of these differences on the number of true positives and true negatives are illustrated in Figure 3. The two models also show differences in their calibration plots (Supplementary Figure 2). In addition, the difference in number of correctly classified individuals between the two models are given. These are calculated assuming, arbitrarily, a 10-year incidence rate of 33%, for example, in a scenario where high-risk individuals have been selected for screening.

**Figure 3.**
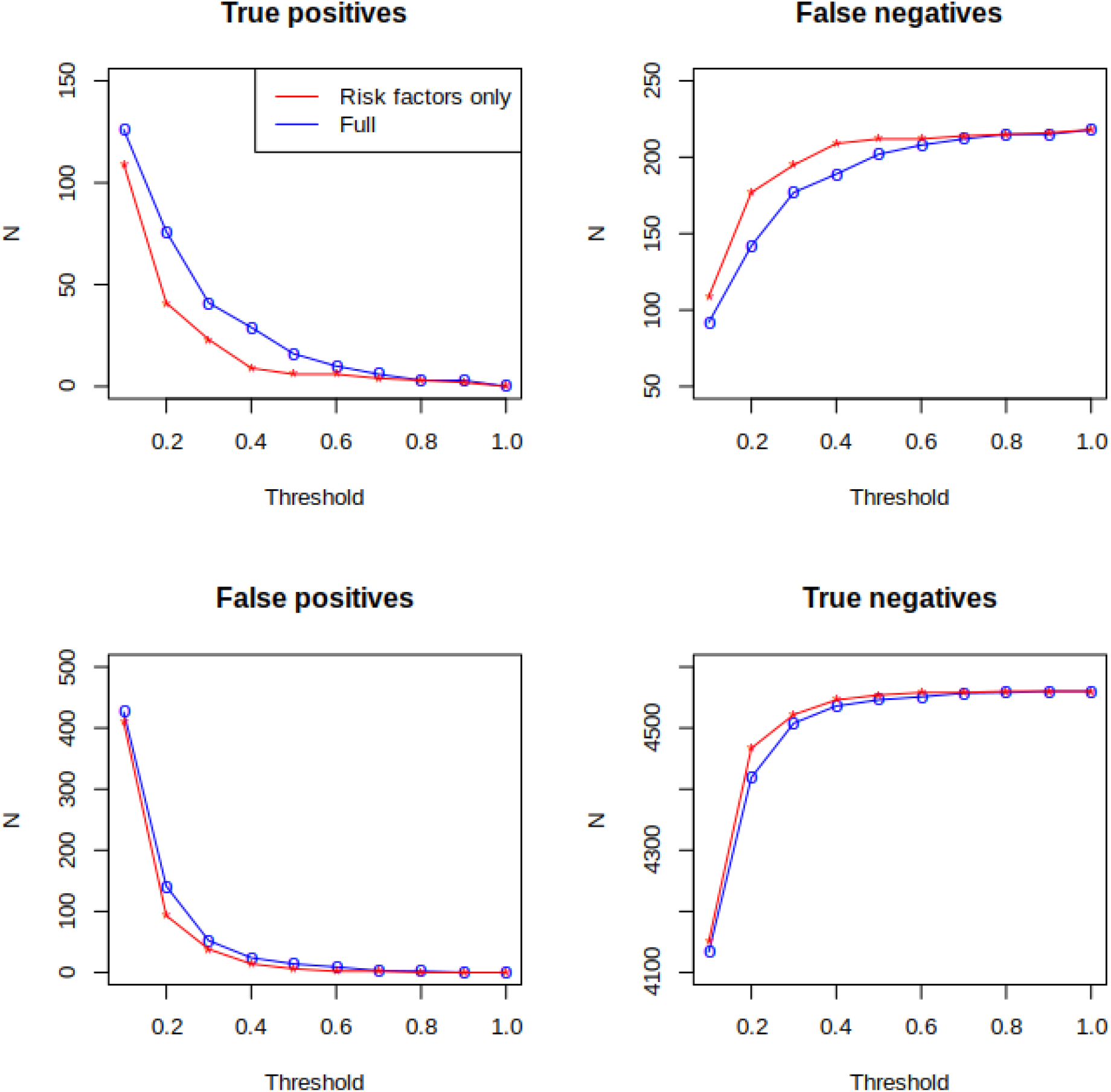
Confusion matrix plot of true/false positives/negatives for the risk factors only and full model in the Generation Scotland test dataset. (Full model uses direct and composite protein EpiScores from Cox PH Lasso.)

### Selected CpGs

The Cox PH lasso model assigned non-zero coefficients to 145 CpGs (**Supplementary Table 7**). After filtering the EWAS Catalog by p-value (p<3.6×10^-8^) [31] and sample size (n>1,000), 119 (82%) of the model-selected CpGs were present. These CpGs corresponded to 742 entries and showed epigenome-wide associations with traits including: serum HDL cholesterol, serum triglycerides, smoking, C-reactive protein, BMI and age (**Supplementary Table 8**).

### Selected Protein EpiScores

The composite protein EpiScore Cox PH lasso model assigned non-zero coefficients to 46 protein EpiScores. Details on the corresponding proteins and genes are given in **Supplementary Table 9**. Out of the selected protein EpiScores, 21 have previously shown associations with incident T2D [9].

### Validation in KORA S4

Prediction of incident diabetes in the KORA S4 cohort using the Cox PH lasso model showed good replication of direct EpiScore performance (p=1.6×10^-5^) with increases of 1.6% and in absolute terms above the null model values for both AUC and PRAUC. Further details are provided in **Supplementary Table 10**.

### EpiScore Prediction of Ongoing Symptomatic COVID-19/Hospitalisation

In all models, incident T2D was predictive of hospitalisation with COVID-19 infection. However, neither the composite-protein nor the direct EpiScore were predictive of the same outcome (**Supplementary Table 11**). Additionally, neither the EpiScores nor incident T2D were predictive of ongoing symptomatic COVID after COVID-19 infection.

## Discussion

Utilising a large cohort with genome-wide epigenetic data and health records linkage to longitudinal primary and secondary care, we have shown that DNAm-based predictors augment standard risk factors in the prediction of incident type 2 diabetes. The best model with traditional risk factors yielded an AUC of 0.839 compared to 0.872 when DNAm was also considered and the PRAUC increased from 0.227 to 0.305. Using a variety of linear and non-linear survival models, we showed that overall, the Cox PH lasso model produced the most predictive direct EpiScore. A composite protein EpiScore also notably increased predictive performance. The direct EpiScore also showed good external validation performance in the KORA S4 cohort. Beyond the T2D analysis presented here, we have developed the *MethylPipeR* R package, along with a user-interface *MethylPipeR-UI*, to facilitate reproducible machine learning time-to-event and binary prediction using DNAm or other types of high-dimensional omics data.

Determining a ‘best’ model is complicated and depends on the trade-off that a user wishes to make. Here, we optimised AUC and PRAUC but binary classification metrics vary by method and/or classification threshold. When using classifiers in clinical settings, decisions need to be made about the number of patients that can be recommended for intervention as well as the acceptable proportion of false positives and false negatives. We showed an increase in correct identification of positives/negatives at varying probability thresholds when adding direct and composite EpiScores above standard risk factors. For instance, given an (arbitrary) incidence rate of 33% (commonly used as a cut-off for high-risk of T2D) [35] over 10 years in a sample of 10,000 individuals, our best model would correctly classify an additional 448 individuals compared to the risk factors only model at a threshold of 0.2 (**Supplementary Table 6**). Given the costs of treating T2D-related complications, our study gives evidence for possible benefits of EpiScores on public health that could also alleviate the financial burden to the NHS. In addition, an assessment of calibration is also critical [36]. Investigation of these related criteria could assist in deciding an optimal threshold given clinical constraints and provide a more comprehensive assessment of model predictions than AUCs or metrics at the commonly-utilised threshold of 0.5.

Several CpGs from the direct EpiScore were previously identified as epigenome-wide significant correlates of traits commonly linked to T2D [14, 17, 37–41]. Future work could investigate overlap between these and time-to-event EWAS studies. Further studies could also include DNAm predictors for traditional risk factors that are included in the null model, such as BMI [8].

Limitations include the relatively small number of disease cases in the dataset, the limited hyperparameter optimisation performed for sBART and the relatively simple variable pre-selection method for tree-ensemble methods. Given the lower performance of these methods compared to the best models in this study, there is potential for additional improvement in predictive performance with further investigation of more advanced feature (pre-)selection. This is particularly important when we consider that the pre-selection step utilised linear models prior to the non-linear model fitting. The model fitting and pre-selection were also performed using the same training set which may have introduced issues associated to post-selection inference [42, 43]. In addition, factors such as overfitting, related individuals in the test set and batch effects between the three rounds of DNAm data processing may all have an effect on test-set AUC. Finally, a small proportion of the linkage codes used to define diabetes included broad terms that were non-specific to T2D; however, late age of onset in these individuals meant there was a high likelihood that they had developed T2D. EpiScores for T2D-associated proteins have also been shown to replicate incident T2D-protein associations within this sample [9] suggesting that the case definitions we use capture biological signals relevant to T2D.

There are numerous strengths to our study. Firstly, the models used capture relationships between CpGs as well as time-to-event information, which is not possible using traditional EWAS methods. Secondly, data linkage to health care measures provided comprehensive T2D incidence data in a very large cohort study, Generation Scotland. Validation performance in the KORA cohort also strengthened evidence for the applicability of the models to other populations. Finally, the R package, *MethylPipeR*, encourages reproducibility and allows others to develop similar predictors on new data with minimal setup.

In conclusion, we have demonstrated the potential for DNA methylation data to provide notable improvement in predictive performance for incident T2D, as compared to traditional risk factors (age, sex, BMI, hypertension, and family history). We evaluated different models with a systematic approach and presented a framework with the ability to generalise to other traits and datasets for training and testing predictors in future studies.

## Supporting information

Supplementary File 1 - TRIPOD checklist

## Data Availability

According to the terms of consent for Generation Scotland participants, access to data must be reviewed by the Generation Scotland Access Committee. Applications should be made to.
All code is available with open access at the following Gitlab repository: https://github.com/marioni-group
MethylPipeR (version 1.0.0) is available at: https://github.com/marioni-group/MethylPipeR
MethylPipeR-UI is available at: https://github.com/marioni-group/MethylPipeR-UI
The informed consents given by KORA study participants do not cover data posting in public databases. However, data are available upon request from KORA Project Application Self-Service Tool (https://epi.helmholtz-muenchen.de/). Data requests can be submitted online and are subject to approval by the KORA Board.

https://github.com/marioni-group

## Declarations

### Ethics approval and consent to participate

All components of Generation Scotland received ethical approval from the NHS Tayside Committee on Medical Research Ethics (REC Reference Number: 05/S1401/89). Generation Scotland has also been granted Research Tissue Bank status by the East of Scotland Research Ethics Service (REC Reference Number: 20-ES-0021), providing generic ethical approval for a wide range of uses within medical research.

The KORA studies were approved by the Ethics Committee of the Bavarian Medical Association (Bayerische Landesärztekammer; S4: #99186) and were conducted according to the principles expressed in the Declaration of Helsinki. All study participants gave their written informed consent.

### Availability of Data and Material

According to the terms of consent for Generation Scotland participants, access to data must be reviewed by the Generation Scotland Access Committee. Applications should be made to.

All code is available with open access at the following Gitlab repository: https://github.com/marioni-group

*MethylPipeR* (version 1.0.0) is available at: https://github.com/marioni-group/MethylPipeR

*MethylPipeR-UI* is available at: https://github.com/marioni-group/MethylPipeR-UI

The informed consents given by KORA study participants do not cover data posting in public databases. However, data are available upon request from KORA Project Application Self-Service Tool (https://epi.helmholtz-muenchen.de/). Data requests can be submitted online and are subject to approval by the KORA Board.

## Acknowledgements

**This research was funded in whole, or in part, by the Wellcome Trust [104036/Z/14/Z, 108890/Z/15/Z, 216767/Z/19/Z]. For the purpose of open access, the author has applied a CC BY public copyright licence to any Author Accepted Manuscript version arising from this submission.**

Generation Scotland received core support from the Chief Scientist Office of the Scottish Government Health Directorates (CZD/16/6) and the Scottish Funding Council (HR03006) and is currently supported by the Wellcome Trust (216767/Z/19/Z). DNA methylation profiling of the Generation Scotland samples was carried out by the Genetics Core Laboratory at the Edinburgh Clinical Research Facility, Edinburgh, Scotland and was funded by the Medical Research Council UK and the Wellcome Trust (Wellcome Trust Strategic Award “STratifying Resilience and Depression Longitudinally” (STRADL; Reference 104036/Z/14/Z). The DNA methylation data assayed for Generation Scotland was partially funded by a 2018 NARSAD Young Investigator Grant from the Brain & Behavior Research Foundation (Ref: 27404; awardee: Dr David M Howard) and by a JMAS SIM fellowship from the Royal College of Physicians of Edinburgh (Awardee: Dr Heather C Whalley). Y.C. is supported by the University of Edinburgh and University of Helsinki joint PhD program in Human Genomics. D.A.G. is supported by funding from the Wellcome Trust 4-year PhD in Translational Neuroscience–training the next generation of basic neuroscientists to embrace clinical research [108890/Z/15/Z]. C.A.V. is a Chancellor’s Fellow funded by the University of Edinburgh. D.L.Mc.C. and R.E.M. are supported by Alzheimer’s Research UK major project grant ARUK-PG2017B−10. R.E.M. is supported by Alzheimer’s Society major project grant AS-PG-19b-010.

Recruitment to the CovidLife study was facilitated by SHARE-the Scottish Health Research Register and Biobank.

SHARE is supported by NHS Research Scotland, the Universities of Scotland and the Chief Scientist Office of the Scottish Government.

The KORA study was initiated and financed by the Helmholtz Zentrum München – German Research Center for Environmental Health, which is funded by the German Federal Ministry of Education and Research (BMBF) and by the State of Bavaria. Furthermore, KORA research has been supported within the Munich Center of Health Sciences (MC-Health), Ludwig-Maximilians-Universität, as part of LMUinnovativ and is supported by the DZHK (German Centre for Cardiovascular Research). The KORA study is funded by the Bavarian State Ministry of Health and Care through the research project DigiMed Bayern (www.digimed-bayern.de).

## Supplementary Methods

### MethylPipeR and MethylPipeR-UI

The analysis pipeline was implemented via a new R package, *MethylPipeR,* along with accompanying user interface, for systematic and reproducible development of complex trait and incident disease predictors. *MethylPipeR* provides functionality for tasks such as model fitting, prediction and performance evaluation as well as automatic logging of experiments and trained models. This is complemented by *MethylPipeR-UI* which provides an interface to the R package functionality while removing the need to write scripts. While *MethylPipeR* was applied to incident T2D prediction with DNA methylation in our experiments, the package is designed for generalised development of predictive models and is applicable to a wide range of omics data and target traits. *MethylPipeR* and *MethlyPipeR-UI* are publicly available at https://github.com/marioni-group/MethylPipeR and https://github.com/marioni-group/MethylPipeR-UI respectively. Supplementary Figure 3 shows an example from the *MethylPipeR-UI* interface including functionality such as data upload, specification of model options and visualisation of model diagnostics.

### Methylation Risk Scores and Incremental Modelling

Composite protein EpiScore models were fit to the training set using 109 protein EpiScores as features [9]. This trained model was used to create a composite protein EpiScore for each individual in the test set. Similarly, direct predictors were fit to the training set using 200,000 CpGs as features and used to create a direct EpiScore for each test set individual.

The following Cox PH models were then fit to the test set to assess the difference in metrics such as the area under the curve (AUC) and area under the precision-recall curve (PRAUC): a risk factors only model; risk factors + protein EpiScore; risk factors + direct EpiScore and a full model (risk factors + protein EpiScore + direct EpiScore). To obtain 10-year survival probabilities in the test set, the cumulative baseline hazard at t=10 was calculated for each model using the Breslow estimator [44].

### Calculating Predictions

To obtain predictions from the incremental models, the following should be applied.

#### Direct EpiScore

For the Cox lasso model, the direct EpiScore can be obtained by summing over the CpG values weighted by their corresponding coefficients (for all non-zero coefficients), as given in **Supplementary Table 12**.

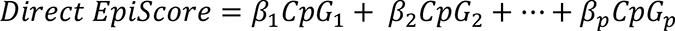

For sBART and RSF, the direct EpiScore requires use of the model object to obtain predictions on new data. These can be found in https://github.com/marioni-group/ and utilised with *MethylPipeR*’s predict function.

#### Composite Protein EpiScore

The composite protein EpiScore can be obtained by summing over the protein EpiScores weighted by their corresponding coefficients (for all non-zero coefficients), as given in **Supplementary Table 12**.

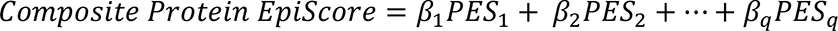

where *q* is the number of protein EpiScores with a non-zero coefficient.

Each protein EpiScore is calculated as the sum over CpG values weighted by their corresponding coefficients (for all non-zero coefficients); the coefficients for each of these can be found in the supplementary materials in Gadd et al. [9]

#### Incremental Cox PH

The linear predictor in an incremental Cox PH model can be calculated by multiplying the relevant EpiScores and risk factors by their corresponding coefficient, given in **Supplementary Table 12**. The cumulative baseline hazard is calculated using the basehaz.gbm function in the gbm R package version 2.1.8 [45]. The 10-year survival probabilities can then be calculated as *S*(*t*) = *exp*[−(*λ*_0_(*t*))]^*exp*(*l*)^at *t* = 10, where *λ*_0_(*t*) is the cumulative baseline hazard and *l* is the linear predictor. The 10-year onset probability is therefore 1 − *S*(10).

### Penalised Cox Proportional-hazards Regression

Since the number of features (200,000) was much greater than the number of individuals in the training set (=9,835 after data preprocessing), a regularisation method was required to reduce overfitting of the Cox PH models.

Cox PH models with lasso penalisation was fit to the training set using glmnet (version 4.1-1) [21, 22] via *MethylPipeR*.

Hyperparameter optimisation and cross validation (CV) were used to select the λ that minimised the partial-likelihood for each Cox PH model corresponding to the minimum partial-likelihood. The training set was divided equally into nine partitions. For each pre-selected value of λ, nine models were fit, each using eight of the partitions as the training set and the ninth for prediction. The mean partial likelihood over the nine models was then calculated. The model using the λ that minimised this was chosen to evaluate on the test set. Nine-fold CV was used to balance the advantages provided by using a greater number of folds with the limitations of the number of cases in each fold as well as required computation time. In addition, individuals belonging to the same family were assigned to the same fold and each contained a similar number of cases to avoid folds with too few cases.

### Random Forest

Random forest [25] is an ensemble machine learning model that estimates a function by averaging the output from a set of independently trained decision trees. During model fitting, each tree is built using a different subset of the variables and the training set to prevent individual trees from overfitting to the whole dataset. In addition, random survival forests adapts the original method to incorporate right-censored time-to-event data [23].

The hyperparameters corresponding to the number of trees (ntrees), the number of variables considered at each tree split (mtry) and the minimum terminal node size (nodesize) were selected using a grid-search CV method. **Supplementary Table 13** shows the set of values that were considered for each hyperparameter. The R package randomForestSRC (version 2.11.0) [27] was used for fitting random survival forests via *MethylPipeR*.

### Bayesian Additive Regression Trees

Bayesian Additive Regression Trees (BART) [26] is a nonparametric method that estimates a function as a sum over a set of regression trees. BART incorporates the ability to model both additive and interaction effects and has shown high predictive performance in comparison with similar methods. To reduce overfitting and model uncertainty in parameters and predictions, BART uses prior distributions over tree-related parameters. Posterior estimates are obtained in a Bayesian framework through Markov Chain Monte Carlo (MCMC).

A variant of BART for survival analysis [24] was used for 10-year onset prediction. 1,000 posterior samples for model parameters were kept after 500 burn-in samples. These were used to generate 10-year survival probabilities on the test set. This resulted in 1,000 survival probabilities for each individual in the test set, the mean of which was used as their survival prediction. MCMC convergence was assessed using Geweke’s diagnostic (gewekediag function in the BART R package). This was calculated on the BART output (yhat.test with t<=10 [24]) for the 1,000 posterior samples using 30 randomly chosen individuals in the test set. Most Z-values (77%) fell within the interval [-1.96, 1.96] suggesting convergence.

Due to the computation time requirements of MCMC sampling and the apparent robustness of BART to hyperparameter misspecification [26], the sBART model was run with hyperparameters set to default. This was performed using the R package BART (version 2.9) [28] via *MethylPipeR*.

### Validation of Best Performing Model in KORA S4

The present analyses are based on a subsample of the participants of the KORA S4 study. KORA (Cooperative Health Research in the Region of Augsburg) is a research platform performing population-based surveys and subsequent follow-ups in the region of Augsburg in Southern Germany [46]. Participants were of German nationality, aged between 25-74 years, 50% female, and sampled from the population registers in the study area where main place of residence was: Ausburg city town, county Ausburg or county Aichach-Friedberg. Each participant completed a health questionnaire, providing details on health status and medication. Blood samples were also taken for assaying of omics data. KORA S4 participants were recruited between 25/10/1999-28/04/2001. This study used a subsample of the 1,451 participants of the KORA S4 study with DNAm and incident T2D data available and no prevalent diabetes at baseline.

The best performing model selected for the Generation Scotland cohort (Cox PH lasso) was used for prediction of incident T2D in the KORA S4 cohort.

For diabetes morbidity, the data are limited to a follow-up of 10 years - starting from KORA S4 recruitment. For incident T2D all prevalent diabetics as well all other diabetes types except T2D cases are excluded. Age, BMI, hypertension, sex as well as self-reported family (mother or father) history of T2D were taken at the baseline of KORA S4. BMI was calculated as the individual’s weight in kg divided by the square of their height in metres.

### EpiScore Prediction of Ongoing Symptomatic COVID-19/Hospitalisation

Self-reported COVID-19 phenotypes were available in a subset of individuals from the Generation Scotland DNA methylation sample who had also participated in the CovidLife surveys (n=2,399) [33]. Ongoing symptomatic COVID-19 phenotypes were ascertained from CovidLife survey 3, (n=1,802 Generation Scotland participants with DNAm profiled), where participants were asked about the total overall time they experienced symptoms in their first/only episode of illness, as well as the whole of their COVID-19 illness. Ongoing symptomatic COVID-19 was defined here as symptoms persisting for at least 4 weeks from infection and is correct as of February 2021, when the survey 3 was administered. Hospitalisation information, derived from the Scottish Morbidity Records (SMR01), was used to obtain COVID-19 hospital admissions using ICD10 codes U07.1 (lab-confirmed COVID-19 diagnosis), and U07.2 (clinically-diagnosed COVID-19). Hospitalisation data is correct as of February 2021. Logistic regression was used to assess the predictive performance of the T2D EpiScores in relation to ongoing symptomatic COVID-19 and severe COVID-19 (i.e. hospitalisation), adjusting for the standard risk factors and incident T2D before a positive COVID-19 test.

---

**Supplementary Figure 1.**
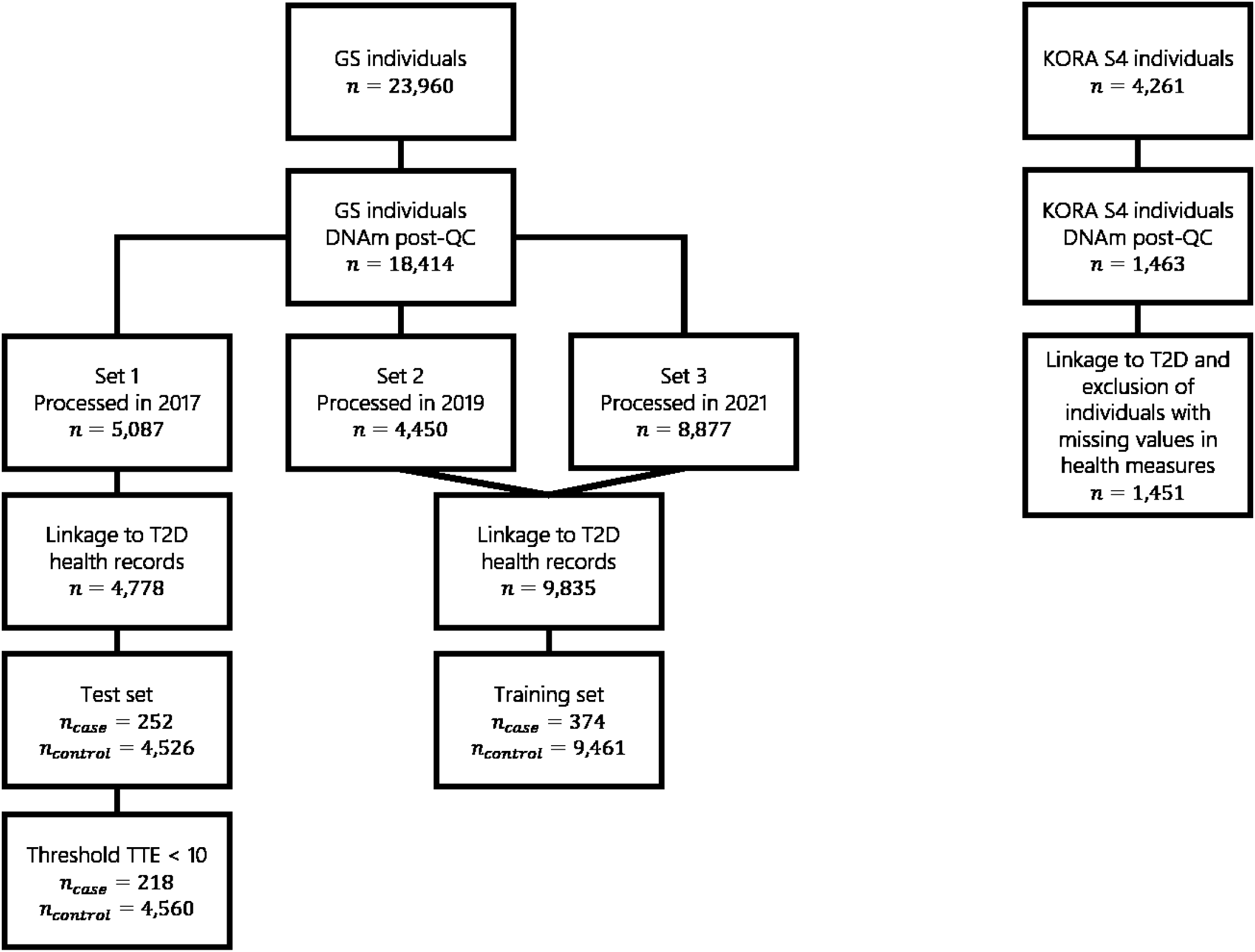
Preprocessing steps for Generation Scotland with number of individuals/cases and controls after each step.

**Supplementary Figure 2.**
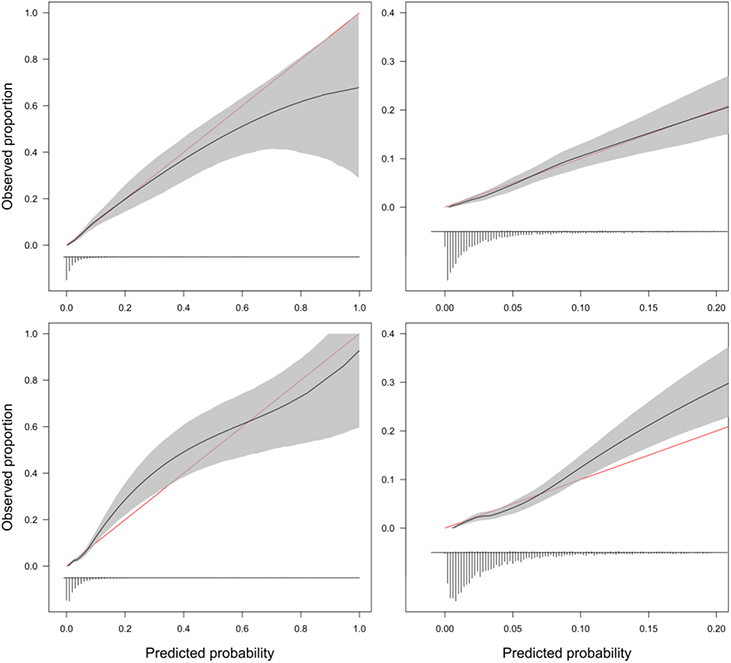
Calibration plots for the full model (risk factors + composite protein EpiScore + Cox PH lasso direct EpiScore) (top-left) and the risk factors only model (bottom-left). The grey area shows 95% confidence intervals calculated from 2000 bootstrap samples. The ideal calibration line (observed = predicted) is shown in red. The histogram shows the distribution of predicted probabilities. The wider confidence intervals at higher predicted probabilities are due to the small number of predictions in those ranges. Most predictions are low in the probability range, emphasised in the zoomed-in plots (top-right and bottom-right).

**Supplementary Figure 3.**
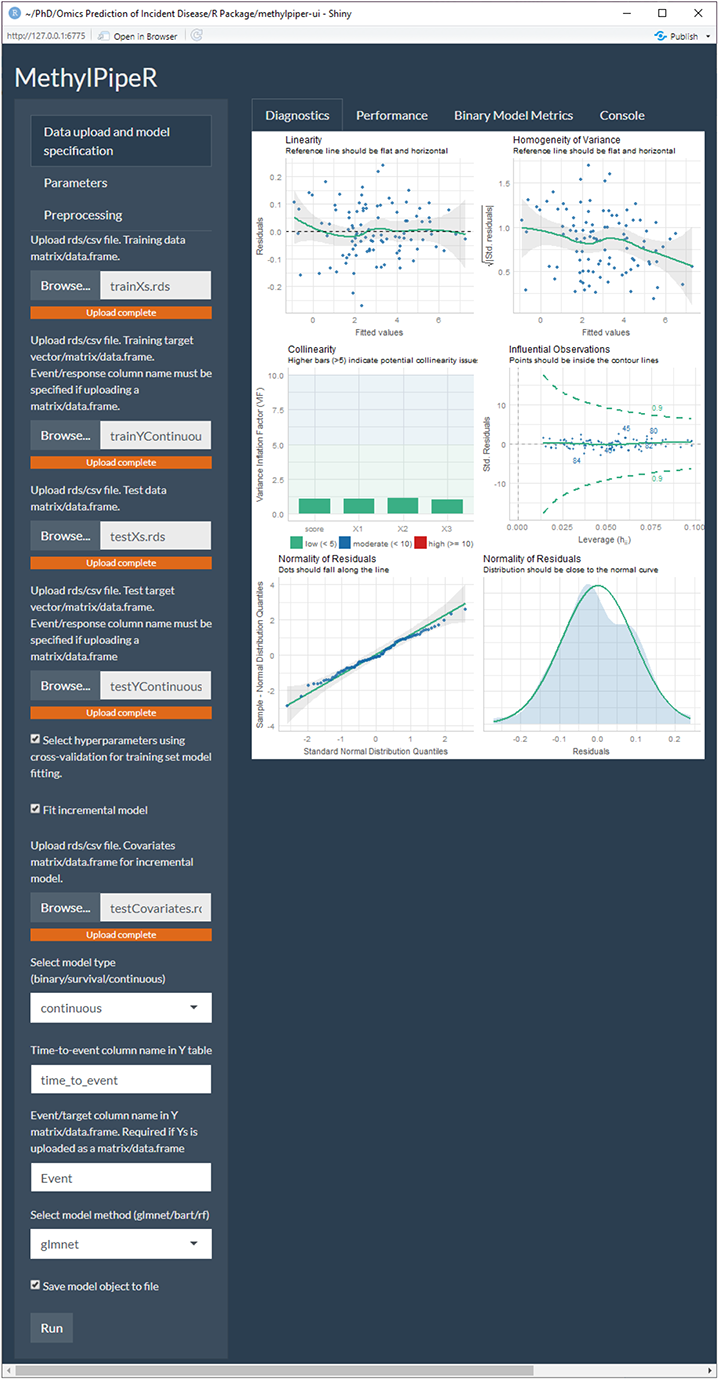
An example from the *MethylPipeR-UI* Shiny app.

## References

1. Saeedi, P., et al., Global and regional diabetes prevalence estimates for 2019 and projections for 2030 and 2045: Results from the International Diabetes Federation Diabetes Atlas. Diabetes research and clinical practice, 2019. 157: p. 107843.

2. Gregg, E.W., N. Sattar, and M.K. Ali, The changing face of diabetes complications. The lancet Diabetes & endocrinology, 2016. 4(6): p. 537–547.

3. Biessels, G.J. and F. Despa, Cognitive decline and dementia in diabetes mellitus: mechanisms and clinical implications. Nature Reviews Endocrinology, 2018. 14(10): p. 591–604.

4. McGurnaghan, S.J., et al., Risks of and risk factors for COVID-19 disease in people with diabetes: a cohort study of the total population of Scotland. The Lancet Diabetes & Endocrinology, 2021. 9(2): p. 82–93.

5. Lee, C.G., et al., Effect of metformin and lifestyle interventions on mortality in the diabetes prevention program and diabetes prevention program outcomes study. Diabetes care, 2021. 44(12): p. 2775–2782.

6. Keng, M.J., et al., Impact of achieving primary care targets in type 2 diabetes on health outcomes and healthcare costs. Diabetes, Obesity and Metabolism, 2019. 21(11): p. 2405–2412.

7. Collins, G.S., et al., Developing risk prediction models for type 2 diabetes: a systematic review of methodology and reporting. BMC medicine, 2011. 9(1): p. 1–14.

8. McCartney, D.L., et al., Epigenetic prediction of complex traits and death. Genome biology, 2018. 19(1): p. 1–11.

9. Gadd, D.A., et al., Epigenetic scores for the circulating proteome as tools for disease prediction. Elife, 2022. 11: p. e71802.

10. Cardona, A., et al., Epigenome-wide association study of incident type 2 diabetes in a British population: EPIC-Norfolk study. Diabetes, 2019. 68(12): p. 2315–2326.

11. Meeks, K.A., et al., Epigenome-wide association study in whole blood on type 2 diabetes among sub-Saharan African individuals: findings from the RODAM study. International journal of epidemiology, 2019. 48(1): p. 58–70.

12. Walaszczyk, E., et al., DNA methylation markers associated with type 2 diabetes, fasting glucose and HbA 1c levels: a systematic review and replication in a case–control sample of the Lifelines study. Diabetologia, 2018. 61(2): p. 354–368.

13. Al Muftah, W.A., et al., Epigenetic associations of type 2 diabetes and BMI in an Arab population. Clinical epigenetics, 2016. 8(1): p. 1–10.

14. Chambers, J.C., et al., Epigenome-wide association of DNA methylation markers in peripheral blood from Indian Asians and Europeans with incident type 2 diabetes: a nested case-control study. The lancet Diabetes & endocrinology, 2015. 3(7): p. 526–534.

15. Nakatochi, M., et al., Epigenome-wide association of myocardial infarction with DNA methylation sites at loci related to cardiovascular disease. Clinical epigenetics, 2017. 9(1): p. 1–9.

16. Wang, X., et al., An epigenome-wide study of obesity in African American youth and young adults: novel findings, replication in neutrophils, and relationship with gene expression. Clinical epigenetics, 2018. 10(1): p. 1–9.

17. Wahl, S., et al., Epigenome-wide association study of body mass index, and the adverse outcomes of adiposity. Nature, 2017. 541(7635): p. 81–86.

18. Haw, J.S., et al., Long-term sustainability of diabetes prevention approaches: a systematic review and meta-analysis of randomized clinical trials. JAMA internal medicine, 2017. 177(12): p. 1808–1817.

19. Collins, G.S., et al., Transparent reporting of a multivariable prediction model for individual prognosis or diagnosis (TRIPOD): the TRIPOD statement. Journal of British Surgery, 2015. 102(3): p. 148–158.

20. Smith, B.H., et al., Cohort Profile: Generation Scotland: Scottish Family Health Study (GS: SFHS). The study, its participants and their potential for genetic research on health and illness. International journal of epidemiology, 2013. 42(3): p. 689–700.

21. Simon, N., et al., Regularization paths for Cox’s proportional hazards model via coordinate descent. Journal of statistical software, 2011. 39(5): p. 1.

22. Friedman, J., T. Hastie, and R. Tibshirani, Regularization paths for generalized linear models via coordinate descent. Journal of statistical software, 2010. 33(1): p. 1.

23. Ishwaran, H., et al., Random survival forests. Annals of Applied Statistics, 2008. 2(3): p. 841–860.

24. Sparapani, R.A., et al., Nonparametric survival analysis using Bayesian additive regression trees (BART). Statistics in medicine, 2016. 35(16): p. 2741–2753.

25. Breiman, L., Random forests. Machine learning, 2001. 45(1): p. 5–32.

26. Chipman, H.A., E.I. George, and R.E. McCulloch, BART: Bayesian additive regression trees. The Annals of Applied Statistics, 2010. 4(1): p. 266–298.

27. Ishwaran, H. and U. Kogalur, Fast unified random forests for survival, regression, and classification (RF-SRC). R package version, 2019. 2(1).

28. Sparapani, R., C. Spanbauer, and R. McCulloch, Nonparametric machine learning and efficient computation with bayesian additive regression trees: the BART R package. Journal of Statistical Software, 2021. 97(1): p. 1–66.

29. Saito, T. and M. Rehmsmeier, The precision-recall plot is more informative than the ROC plot when evaluating binary classifiers on imbalanced datasets. PloS one, 2015. 10(3): p. e0118432.

30. Battram, T., et al., The EWAS Catalog: a database of epigenome-wide association studies. 2021.

31. Saffari, A., et al., Estimation of a significance threshold for epigenome-wide association studies. Genetic epidemiology, 2018. 42(1): p. 20–33.

32. Wichmann, H.-E., et al., KORA-gen-resource for population genetics, controls and a broad spectrum of disease phenotypes. Das Gesundheitswesen, 2005. 67(S 01): p. 26–30.

33. Fawns-Ritchie, C., et al., CovidLife: a resource to understand mental health, well-being and behaviour during the COVID-19 pandemic in the UK. Wellcome Open Research, 2021. 6(176): p. 176.

34. Shah, W., et al., Managing the long term effects of covid-19: summary of NICE, SIGN, and RCGP rapid guideline. bmj, 2021. 372.

35. Ekoe, J.-M., R. Goldenberg, and P. Katz, Screening for diabetes in adults. Canadian journal of diabetes, 2018. 42: p. S16–S19.

36. Van Calster, B., et al., Calibration: the Achilles heel of predictive analytics. BMC medicine, 2019. 17(1): p. 1–7.

37. Demerath, E.W., et al., Epigenome-wide association study (EWAS) of BMI, BMI change and waist circumference in African American adults identifies multiple replicated loci. Human molecular genetics, 2015. 24(15): p. 4464–4479.

38. Mendelson, M.M., et al., Association of body mass index with DNA methylation and gene expression in blood cells and relations to cardiometabolic disease: a Mendelian randomization approach. PLoS medicine, 2017. 14(1): p. e1002215.

39. Sayols-Baixeras, S., et al., Identification and validation of seven new loci showing differential DNA methylation related to serum lipid profile: an epigenome-wide approach. The REGICOR study. Human molecular genetics, 2016. 25(20): p. 4556–4565.

40. Braun, K.V., et al., Epigenome-wide association study (EWAS) on lipids: the Rotterdam Study. Clinical epigenetics, 2017. 9(1): p. 1–11.

41. Kriebel, J., et al., Association between DNA methylation in whole blood and measures of glucose metabolism: KORA F4 study. PloS one, 2016. 11(3): p. e0152314.

42. Lee, J.D., et al., Exact post-selection inference, with application to the lasso. The Annals of Statistics, 2016. 44(3): p. 907–927.

43. Taylor, J. and R. Tibshirani, Post-selection inference for-penalized likelihood models. Canadian Journal of Statistics, 2018. 46(1): p. 41–61.

44. Lin, D., On the Breslow estimator. Lifetime data analysis, 2007. 13(4): p. 471–480.

45. Ridgeway, G., Generalized Boosted Models: A guide to the gbm package. Update, 2007. 1(1): p. 2007.

46. Holle, R., et al., KORA-a research platform for population based health research. Das Gesundheitswesen, 2005. 67(S 01): p. 19–25.

